# Significant and sustained decrease in non-SARS-CoV-2 respiratory viral infections during COVID-19 public health interventions

**DOI:** 10.1101/2021.05.11.21256147

**Authors:** Jeffrey D. Whitman, Phong Pham, Caryn Bern, Elaine M. Dekker, Barbara L. Haller, Vivek Jain, Lisa G. Winston

## Abstract

Public health interventions to decrease the spread of SARS-CoV-2 were largely implemented in the United States during spring 2020. This study evaluates the additional effects of these interventions on non-SARS-CoV-2 respiratory viral infections from a single healthcare system in the San Francisco Bay Area. The results of a respiratory pathogen multiplex polymerase chain reaction panel intended for inpatient admissions were analyzed by month between 2019 and 2020. We found major decreases in the proportion and diversity of non-SARS-CoV-2 respiratory viral illnesses in all months following masking and shelter-in-place ordinances. These findings suggest real-world effectiveness of nonpharmaceutical interventions on droplet-transmitted respiratory infections.

## INTRODUCTION

Public health interventions to decrease the spread of SARS-CoV-2 have been in place in the San Francisco Bay Area since spring 2020. A shelter-in-place order went into effect in Bay Area counties on March 16, 2020 with dynamic restrictions over the summer, fall and winter of 2020 (1). An order requiring face coverings went into effect April 17, 2020 (2). While these types of nonpharmaceutical interventions have been linked to dramatic reductions in the rates of influenza (3, 4), there has been a paucity of data examining rates of circulation of other common respiratory viruses.

## METHODS

Results of a multiplex polymerase chain reaction respiratory pathogen panel (RPP) assay (FilmArray RP2, Biofire Diagnostics) were extracted from the Zuckerberg San Francisco General Hospital (ZSFG) clinical laboratory information system from January 2019 to December 2020; a period spanning approximately one year before and during the COVID-19 pandemic. The RPP panel tests for 17 viral pathogens and 4 bacterial organisms. At our center, this test is utilized for evaluation of infectious respiratory diseases in patients admitted to the hospital. To assess trends in incidence of respiratory pathogens during the COVID-19 pandemic, we compared the number and percentage of positive results (total and by pathogen) by month. We tested for significance using Chi-square or Fisher exact tests. All statistical analyses were performed in Prism 9.0.0 (GraphPad Software). This project was approved by UCSF Human Research Protections Program (IRB# 20-32769) and ZSFG protocol approval board.

## RESULTS

A total of 1,484 and 2,037 RPP tests were analyzed during 2019 and 2020, respectively. Percent positivity of tests by month ranged from 13.6%-39.1% during 2019 and 2020 pre-COVID-19 interventions (January, 2019-March, 2020) and 0.0%-11.1% during 2020 post-interventions period (April-December, 2020) (Figure 1). The proportion of total positive tests was not significantly different for January, February, and March of 2019 versus the same months in 2020. However, the proportion of positive tests was lower for every month April-December 2020 during COVID-19 restrictions compared to 2019. Human rhinoviruses/enteroviruses (HRE) were the respiratory pathogens most frequently detected across all years (Table 1). During the April-December 2020 post-interventions period, 89.6% (43/48) of positive results were HRE versus 55.0% (177/322) over the same period in 2019. Zero influenza cases were detected during the 2020-2021 influenza season for this time period.

**Table 1.** Respiratory infections identified by multiplex molecular pathogen panel testing from 2019 and 2020.

|  | Month | Total Tested | No. Positive (%) | ADV | CV2 | CVC | CVH | CVNL | FLUA | FLUB | HMP | HRE | MPP | PIV1 | PIV2 | PIV3 | PIV4 | RSV |
| --- | --- | --- | --- | --- | --- | --- | --- | --- | --- | --- | --- | --- | --- | --- | --- | --- | --- | --- |
| Pre-COVID-19 Interventions | Jan 2019 | 117 | 37 (31.6) | 0 | 2 | 5 | 0 | 3 | 2 | 0 | 1 | 16 | 0 | 0 | 3 | 0 | 2 | 3 |
|  | Feb 2019 | 152 | 46 (30.3) | 1 | 0 | 11 | 0 | 2 | 4 | 0 | 5 | 17 | 0 | 0 | 1 | 2 | 0 | 3 |
|  | Mar 2019 | 135 | 40 (29.6) | 0 | 1 | 6 | 2 | 3 | 9 | 0 | 3 | 13 | 0 | 0 | 0 | 0 | 0 | 3 |
|  | Apr 2019 | 137 | 52 (38.0) | 2 | 0 | 2 | 0 | 2 | 7 | 0 | 10 | 17 | 0 | 0 | 0 | 6 | 0 | 6 |
|  | May 2019 | 138 | 52 (37.7) | 1 | 0 | 3 | 0 | 0 | 1 | 0 | 6 | 26 | 0 | 0 | 0 | 13 | 0 | 2 |
|  | Jun 2019 | 108 | 24 (22.2) | 0 | 0 | 0 | 2 | 0 | 1 | 0 | 1 | 17 | 0 | 0 | 0 | 3 | 0 | 0 |
|  | Jul 2019 | 81 | 11 (13.6) | 0 | 0 | 0 | 0 | 0 | 1 | 0 | 0 | 4 | 2 | 1 | 0 | 3 | 0 | 0 |
|  | Aug 2019 | 75 | 22 (29.3) | 1 | 0 | 0 | 0 | 0 | 0 | 0 | 0 | 18 | 1 | 0 | 0 | 1 | 1 | 0 |
|  | Sep 2019 | 110 | 34 (30.9) | 1 | 0 | 0 | 0 | 1 | 0 | 0 | 0 | 29 | 1 | 0 | 1 | 0 | 0 | 1 |
|  | Oct 2019 | 126 | 37 (29.4) | 2 | 0 | 1 | 0 | 0 | 1 | 0 | 1 | 27 | 1 | 1 | 0 | 1 | 0 | 2 |
|  | Nov 2019 | 121 | 31 (25.6) | 0 | 0 | 3 | 2 | 1 | 4 | 1 | 1 | 9 | 0 | 3 | 1 | 0 | 0 | 6 |
|  | Dec 2019 | 184 | 59 (32.1) | 0 | 0 | 3 | 8 | 1 | 6 | 1 | 3 | 30 | 0 | 3 | 1 | 0 | 0 | 3 |
|  | Jan 2020 | 215 | 84 (39.1) | 2 | 4 | 0 | 3 | 0 | 15 | 1 | 13 | 39 | 2 | 1 | 0 | 0 | 0 | 4 |
|  | Feb 2020 | 222 | 79 (35.6) | 2 | 5 | 3 | 11 | 2 | 11 | 1 | 8 | 27 | 0 | 0 | 0 | 0 | 0 | 9 |
|  | Mar 2020 | 685 | 203 (29.6) | 13 | 8 | 3 | 14 | 3 | 28 | 2 | 24 | 81 | 3 | 2 | 0 | 1 | 1 | 20 |
| Post-COVID-19 Interventions | Apr 2020 | 156 | 5 (3.2) | 0 | 0 | 0 | 0 | 0 | 0 | 0 | 0 | 4 | 0 | 0 | 0 | 0 | 0 | 1 |
|  | May 2020 | 118 | 1 (3.2) | 1 | 0 | 0 | 0 | 0 | 0 | 0 | 0 | 0 | 0 | 0 | 0 | 0 | 0 | 0 |
|  | Jun 2020 | 68 | 2 (2.9) | 0 | 0 | 0 | 0 | 1 | 0 | 0 | 0 | 1 | 0 | 0 | 0 | 0 | 0 | 0 |
|  | Jul 2020 | 53 | 2 (3.8) | 0 | 0 | 0 | 0 | 0 | 0 | 0 | 0 | 1 | 0 | 0 | 0 | 0 | 1 | 0 |
|  | Aug 2020 | 69 | 0 (0.0) | 0 | 0 | 0 | 0 | 0 | 0 | 0 | 0 | 0 | 0 | 0 | 0 | 0 | 0 | 0 |
|  | Sep 2020 | 53 | 1 (1.9) | 0 | 0 | 0 | 0 | 0 | 0 | 0 | 0 | 1 | 0 | 0 | 0 | 0 | 0 | 0 |
|  | Oct 2020 | 54 | 6 (11.1) | 0 | 0 | 0 | 0 | 0 | 0 | 0 | 0 | 6 | 0 | 0 | 0 | 0 | 0 | 0 |
|  | Nov 2020 | 109 | 11 (10.1) | 0 | 0 | 0 | 0 | 0 | 0 | 0 | 0 | 11 | 0 | 0 | 0 | 0 | 0 | 0 |
|  | Dec 2020 | 235 | 20 (8.5) | 1 | 0 | 0 | 0 | 0 | 0 | 0 | 0 | 19 | 0 | 0 | 0 | 0 | 0 | 0 |

**Abbreviations:** ADV, adenovirus; CV2, coronavirus 229E; CVC, coronavirus OC43; CVH, coronavirus HKU1; CVNL, coronavirus NL63; FLUA, influenza A virus (including A/H3 and A/H1 2009); FLUB, influenza B virus; HMP, human metapneumovirus; HRE, human rhino/enterovirus; MPP, *Mycoplasma pneumoniae*; PIV1, parainfluenza type 1; PIV3, parainfluenza type 3; PIV4 parainfluenza, type 4; RSV, respiratory syncytial virus.

**Figure 1.**
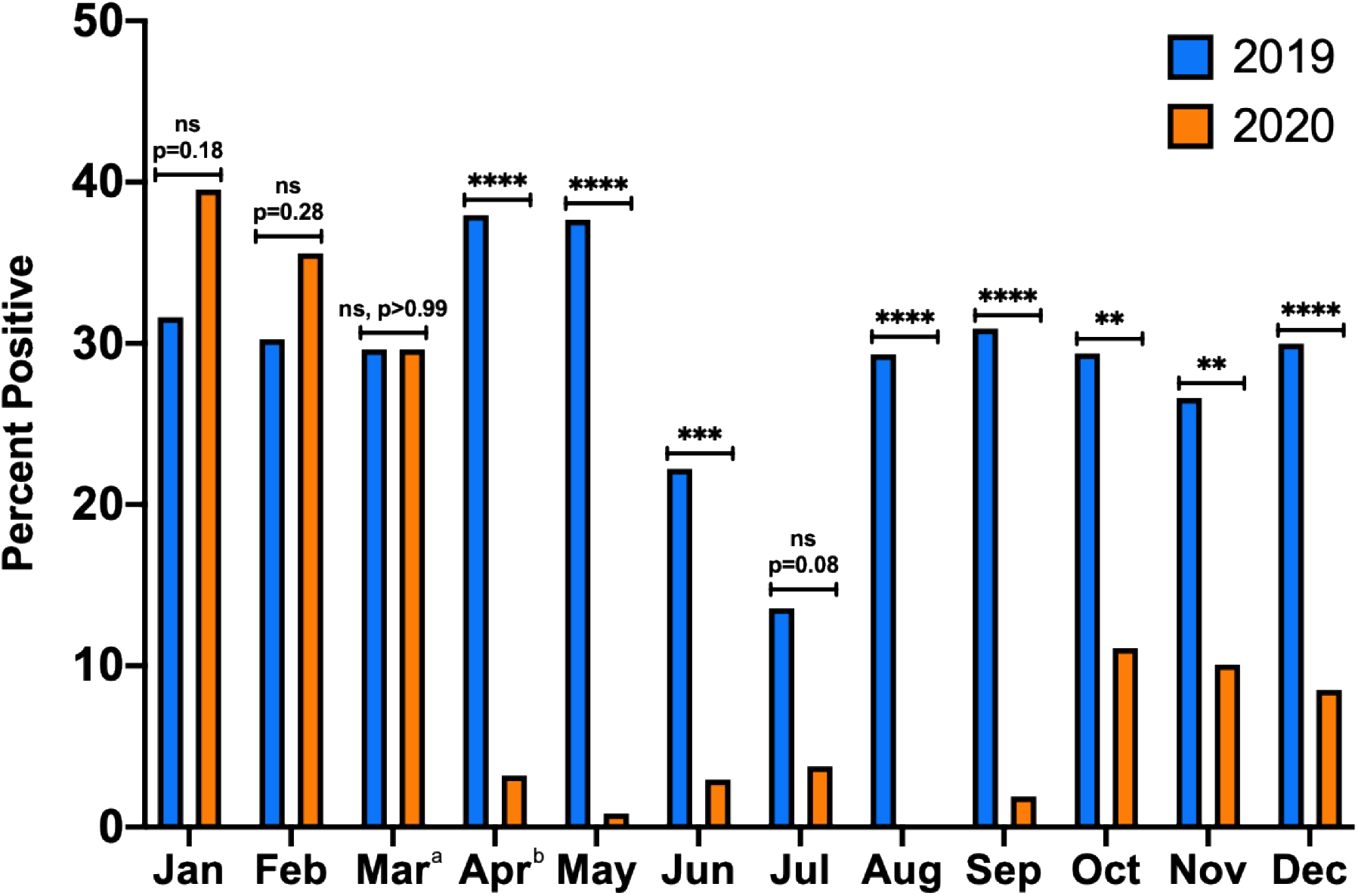
Proportion of positive respiratory pathogen tests by month in 2019 compared to 2020. Proportion of tests positive in 2019 and 2020 are shown by month. Respiratory viral illness diagnoses became significantly less frequent from March to April 2020, and from April-December 2020, the proportion of tests positive was consistently lower compared to the corresponding month in 2019. NS, not significant; *, P<0.05; **, P<0.01; ***, P<0.001; ****, p<0.0001. ^a^ Shelter-in-Place went into effect on March 16, 2020. ^b^ Masking ordinance went into effect on April 17, 2020.

## DISCUSSION

The COVID-19 pandemic began affecting the United States in March 2020 (5). To mitigate disease transmission in the San Francisco Bay Area, public health orders to shelter-in-place and wear face coverings were imposed and have largely been followed. The effectiveness of these interventions has primarily been measured in relation to SARS-CoV-2 incident infections, for which testing availability and indications and public health mitigation strategies have varied widely. The effect of mitigation measures on non-COVID-19 respiratory infections has mainly been seen in the profound reduction of influenza (3, 4). However, the seasonal nature of influenza has prevented generalization of this trend to other respiratory viruses, particularly those that circulate year-round. We observed a striking decrease in the number and percentage of all respiratory viruses after public health interventions began in March 2020 and observed a sustained low rate of respiratory virus detection throughout the summer, fall and early winter of 2020.

Our study contains certain limitations. We employed a cross-sectional design comparing two years of data from a single healthcare system. As such, we cannot control for year-to-year variations in viral respiratory illnesses or demographic confounders in our study population. Our data may also be affected by reduction of health services or avoidance of medical care directly from COVID-19 and evolving local policies regarding allowable indoor businesses and personal gathering (6, 7). Despite this, our focus on inpatients tested for respiratory infections, rather than the larger population of outpatients, should decrease variation in testing practices that may occur from workup of mild and undifferentiated illness.

While our findings add to the evidence regarding effective disease mitigation strategies (8–10), it is also notable that the low rate of respiratory virus detection remained low during intermittent months of increased public interaction and movement in the Bay Area, including periods when businesses were allowed to partially reopen, times at which civil protests occurred, and periods of increased summer outdoor recreational activities. This suggests a central role for masking and reduction of enclosed space gathering for the disruption of respiratory viral transmission.

Since our study population focuses on prospective inpatient admissions, these results primarily represent moderate and severe infections requiring high-level care. As such, these findings also suggest that community-based strategies have downstream effects on inpatients with respiratory illness. While we cannot conclude which sequence or combination of nonpharmaceutical interventions is most effective, the sustained decrease in respiratory viral illnesses we observed argues for further investigation of the impact of these strategies, particularly those aiming to reduce droplet transmission, in personal, healthcare and workplace settings.

## Data Availability

All relevant data is available in the tables and figures of this manuscript.

## REFERENCES

1. Department of Public Health City and County of San Francisco. 2020. ORDER OF THE HEALTH OFFICER OF THE CITY AND COUNTY OF SAN FRANCISCO DIRECTING ALL INDIVIDUALS LIVING IN THE COUNTY TO SHELTER AT THEIR PLACE OF RESIDENCE EXCEPT THAT THEY MAY LEAVE TO PROVIDE OR RECEIVE CERTAIN ESSENTIAL SERVICES OR ENGAGE IN CERTAIN ESSENTIAL ACTIVITIES AND WORK FOR ESSENTIAL BUSINESS AND GOVERNMENT SERVICES;. https://sfbos.org/sites/default/files/20200316_DPH_Order_of_the_Health_Officer_C19-07.pdf. Accessed December 1.

2. Department of Public Health City and County of San Francisco. 2020. ORDER OF THE HEALTH OFFICER OF THE CITY AND COUNTY OF SAN FRANCISCO GENERALLY REQUIRING MEMBERS OF THE PUBLIC AND WORKERS TO WEAR FACE COVERINGS https://sfbos.org/sites/default/files/20200417_FINAL_Order_No_C19-12.pdf. Accessed December 1.

3. Olsen SJ, Azziz-Baumgartner E, Budd AP, Brammer L, Sullivan S, Pineda RF, Cohen C, Fry AM. 2020. Decreased Influenza Activity During the COVID-19 Pandemic - United States, Australia, Chile, and South Africa, 2020. MMWR Morb Mortal Wkly Rep 69:1305–1309.

4. Centers for Disease Control and Prevention. Weekly U.S. Influenza Surveillance Report. https://www.cdc.gov/flu/weekly/index.htm. Accessed March 12th.

5. Johns Hopkins University COVID Resource Center. 2020. COVID-19 United States Cases by County, on Johns Hopkins University. https://coronavirus.jhu.edu/us-map. Accessed 11-25-2020.

6. Whaley CM, Pera MF, Cantor J, Chang J, Velasco J, Hagg HK, Sood N, Bravata DM. 2020. Changes in Health Services Use Among Commercially Insured US Populations During the COVID-19 Pandemic. JAMA Netw Open 3:e2024984.

7. Czeisler ME, Marynak K, Clarke KEN, Salah Z, Shakya I, Thierry JM, Ali N, McMillan H, Wiley JF, Weaver MD, Czeisler CA, Rajaratnam SMW, Howard ME. 2020. Delay or Avoidance of Medical Care Because of COVID-19-Related Concerns - United States, June 2020. MMWR Morb Mortal Wkly Rep 69:1250–1257.

8. Hartley DM, Perencevich EN. 2020. Public Health Interventions for COVID-19: Emerging Evidence and Implications for an Evolving Public Health Crisis. JAMA 323:1908–1909.

9. Joo H, Miller GF, Sunshine G, Gakh M, Pike J, Havers FP, Kim L, Weber R, Dugmeoglu S, Watson C, Coronado F. 2021. Decline in COVID-19 Hospitalization Growth Rates Associated with Statewide Mask Mandates - 10 States, March-October 2020. MMWR Morb Mortal Wkly Rep 70:212–216.

10. Guy GP, Jr., Lee FC, Sunshine G, McCord R, Howard-Williams M, Kompaniyets L, Dunphy C, Gakh M, Weber R, Sauber-Schatz E, Omura JD, Massetti GM, Cdc Covid-19 Response Team MPAU, Program CDCPHL. 2021. Association of State-Issued Mask Mandates and Allowing On-Premises Restaurant Dining with County-Level COVID-19 Case and Death Growth Rates - United States, March 1-December 31, 2020. MMWR Morb Mortal Wkly Rep 70:350–354.

